# Estimating population structure using epigenome-wide methylation data

**DOI:** 10.1101/2025.09.01.25334865

**Authors:** Ziqing Wang, Kent D Taylor, Jerome I Rotter, Stephen S Rich, Yinan Zheng, Lifang Hou, Xiuqing Guo, Jan Bressler, Laura M Raffield, Yongmei Liu, Robert Kaplan, Donald M Lloyd-Jones, Alanna C. Morrison, Myriam Fornage, Tamar Sofer, the TOPMed Epigenetics working group

## Abstract

**Introduction:** In epigenome-wide association analysis (EWAS), unaddressed population stratification often leads to inflation. We aimed to compute methylation population scores (MPSs) that predict genetic principal components (GPCs) using a feature selection and regression approach.

**Methods:** We used multi-ethnic methylation data (Illumina 450K/EPIC array) from unrelated MESA (n=929), CARDIA (n=1123), JHS (n=1365), ARIC (n=2338), and HCHS/SOL (n=1475) individuals, randomly assigning 85% of participants from each cohort to a training dataset and the remaining 15% to a test dataset. First, we estimated the associations of GPCs with each available CpG methylation site using linear regression within each cohort, adjusting for age, sex, smoking status, race/ethnic background (as a proxy for background information associated with lifestyle and other environmental exposures that may impact methylation), alcohol use status, body mass index, and cell type proportions. We meta-analyzed the associations across cohorts and selected CpG sites with association FDR-adjusted q-value <0.05. We next aggregated individual-level data across the cohort-specific training datasets, and applied two-stage weighted least squares Lasso regression, with the GPCs as the outcomes and the selected CpG sites as penalized predictors, adjusting for the aforementioned covariates. The developed MPSs are the weighted sum of selected CpG sites from the Lasso. To evaluate the developed MPSs, we constructed them in the test dataset, and compared them with GPCs, and with MPSs constructed based on a previously-published paper. Comparison was based on correlation analysis and data visualization. We demonstrate the use of the MPSs in EWAS.

**Results:** In the test dataset, the MPSs were highly correlated with GPCs, with correlation decreasing, though not monotonically, for later components. Specifically, MPS1 and GPC1 had R2= 0.99, while MPS7 and GPC7 had R2=0.27 (the lowest observed correlation). In data visualization, MPSs had similar patterns as GPCs in differentiating self-reported White, Black, and Hispanic/Latino groups, while outperforming MPC constructed using alternative published methods. MPSs showed comparable performance to GPCs in reducing some of the inflation in EWAS.

**Conclusions:** Methylation-based population scores provide a reliable estimate of population structure in the data and can complement GPCs when genetic data are absent. Unlike previous methods based on unsupervised methylation PCA, MPSs uses supervised learning with covariate adjustment to capture genetic structure across diverse populations. The weights for each GPCs derived in our study can be applied to generate MPSs in other studies.

## Introduction

Epigenome-wide association studies (EWAS) and genome wide association studies (GWAS) are powerful approaches to identify epigenetic and genetic associations, respectively, with complex traits and diseases. In both types of analyses, it is essential to address population stratification, a form of bias caused by linkage disequilibrium and allele frequency divergence between subpopulations, which arise from populations’ evolutionary and demographic histories^1,2^. Failure to account for population structure can lead to test statistic inflation and increased false positive findings, thus complicating the identification of true biological signals. The most common method to address population stratification is to include genetic principal components (GPCs), derived from principal components analysis (PCA) of genome-wide genetic data, as covariates in the association analysis. Consequently, the absence or incompleteness of genetic data often limits the sample size and thereby reduces the statistical power of EWAS association analyses.

DNA methylation (DNAm) is an epigenetic modification that can influence phenotype and disease outcomes by regulating gene expression through transcription and chromosomal inactivation^3,4^. Notably, DNAm is shaped by both genetic factors and environmental exposures^5,6^, ranging from lifestyle and socioeconomic stressors^7^ to infectious agents^8^, and is characterized by its long-term stability, with potential transgenerational impact^9^. That said, previous studies have attempted to compute methylation-based PCs by PCA on genome-wide DNA methylation data or using CpG sites highly correlated with single nucleotide polymorphisms (SNPs) to capture population structure^8,10,11^. For instance, Rahmani and colleagues proposed to construct methylation PC (MPC) by applying PCA on a set of genetically informative CpGs in close vicinity to SNPs selected by linear model^10^. These findings demonstrate that genetic ancestry information is also embedded in one’s DNAm profiles, offering a promising opportunity for addressing population structure in epigenome-wide association studies when genetic data are unavailable.

One potential challenge with using PCs obtained by PCA of genome-wide methylation rather than genetic data, is that they may inadvertently capture technical or demographic variation, such as differences in cell type composition or age, since DNA methylation is influenced by a wide range of factors beyond genetic ancestry^11,12^. In this study, we develop methylation-based measures of GPCs, which we term “Methylation population scores (MPS)”. The MPSs are methylation scores that predict GPCs. We develop them using a feature selection approach informed by GPCs, while accounting for additional confounding variables, including age, sex, estimated cell type proportions, and environmental factors, including geographical locations and key lifestyles. We use methylation and genetic data from five multi-ethnic cohorts participating in the Trans-Omics in Precision Medicine (TOPMed) program, encompassing individuals self-reporting of White, Black, Chinese, and Hispanic backgrounds. We further compared our MPSs to MPC computed through PCA using reference list provided by Rahmani et al^10^. The significance of this study is two-fold, 1) develop MPSs based on available, continuous, genetic ancestry information (in the form of genetic PCs), meanwhile controlling for environmental and technical confounders through covariate adjustment; and 2) incorporate a broader range of population groups and a larger sample size compared to previous studies, which primarily focused on individuals of White and Black backgrounds.

## Methods

Figure 1 provides an overview of the study. We used methylation data (Illumina 850K/EPIC array in primary analysis, limiting to CpG sites available on the Illumina HumanMethylation450 BeadChip in secondary analysis, from genetically unrelated, individuals from 5 studies representing multiple race/ethnicity backgrounds from the TOPMed program: Multi-Ethnic Study of Atherosclerosis (MESA), Coronary Artery Risk Development in Young Adults (CARDIA), Jackson Heart Study (JHS), Atherosclerosis Risk in Communities (ARIC) study, and the Hispanic Community Health Study/Study of Latinos (HCHS/SOL). Within each cohort, we randomly assigned 85% of participants to a training dataset and the remaining to a test dataset. We developed MPSs using CpG sites identified by feature selection methods trained on GPCs, and performed multiple analyses to evaluate their performance.

**Figure 1.**
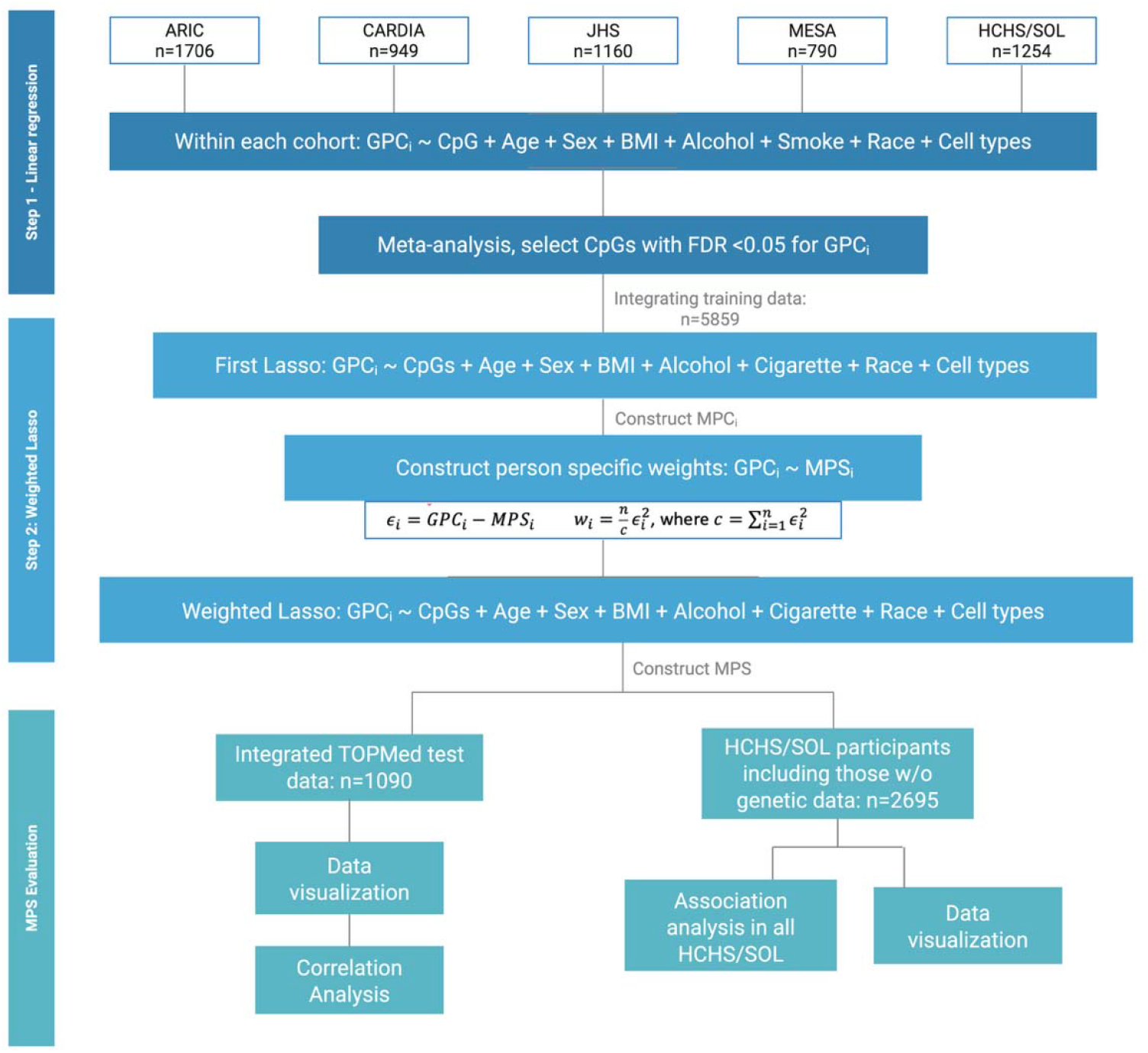
Analysis plan of the study. ARIC: Atherosclerosis Risk in Communities study; CARDIA: Coronary Artery Risk Development in Young Adults; JHS: Jackson Heart Study; MESA: Multi-Ethnic Study of Atherosclerosis; HCHS/SOL: Hispanic Community Health Study/Study of Latinos

### The Trans-Omics for Precision Medicine (TOPMed) program

The TOPMed program aims to improve diagnosis, treatment and prevention of heart, lung, blood and sleep disorders by elucidating genetic and phenotypic data over 130,000 participants from more than 80 participating studies^13^, available through dbGaP (Database of Genotypes and Phenotypes)^14^. To minimize batch bias across cohorts, sequencing centers and experiment years, TOPMed standardized laboratory methods to a single pipeline and performed variant and genotype calling jointly on all samples in a given TOPMed freeze^13^. For this study, we obtained TOPMed whole genome sequencing (WGS) data from the Freeze 10 release.

### Whole Genome Sequencing (WGS)

DNA samples were extracted from whole blood samples collected from each participating cohort, before being processed for pair-ended 150 bp sequencing with a mean depth of 30x using Illumina HiSeq X Ten instruments at TOPMed sequencing centers^14^. As aforementioned, generated reads were first mapped to the human-genome build GRCh38 following a common pipeline across all sequencing centers. Variant discovery and genotyping were performed jointly across all available samples in freeze 10 using GotCloud pipeline^15^. Quality control included variant inclusion by minor allele frequency (MAF > 0.01), genotypes with a minimal depth of more than 10x, and sample-level checks for pedigree errors, self-reported and genetic sex discrepancies, and discordance with previous genotyping array data^14^. Genetic principal components (GPCs) and genetic relatedness were also centrally computed using the PC-AiR and PC-Relate algorithms^16^. Among our study participants, a set of unrelated individuals was defined as a maximal set of individuals where twice the kinship coefficient was lower than 0.0625*2 for all pairs of individuals, limiting relatedness to 3^rd^ degree.

### Epigenome-wide methylation quantification

Methylation profiling for ARIC, CARDIA, and HCHS/SOL was performed at the University of Washington Northwest Genomics Center (NWGC), where DNA samples underwent normalization and bisulfite conversion using the EZ-96 DNA Methylation kit (#D5003; ZYMO Research). Methylation data for “NHLBI TOPMed: Multi-Ethnic Study of Atherosclerosis (MESA)” was performed at Keck Molecular Genomics Core Facility.” DNAm profiles were generated using the MethylationEPIC BeadChip and quality checked using Illumina GenomeStudio software (v2.0.3) to include samples with a genotyping call rate of 0.98 or higher. Samples passing quality control were normalized by normal-exponential deconvolution using out-of-band probes (Noob) background subtraction^17^ using the R package Minfi^18^ to obtain beta-values. We estimated cell type subpopulations from the processed methylation data for each cohort using reference-based Houseman’s method^19^.

HCHS/SOL DNAm samples were also profiled using the Infinium MethylationEPIC array at the HCHS/SOL Data Coordinating Center (DCC) at the University of North Carolina at Chapel Hill, and examined for sex and SNPs mismatches, control and blind duplicate, followed by normalization using the R package SeSAMe^20^ to mask 105,454 probes, dye bias correction, and Noob normalization^17,18^. The obtained beta-values were further corrected for type-2 probe bias using BMIQ method from the WateRmelon package^21^, and underwent ComBat batch correction^22^. Subsequently, cell type proportions were also estimated using Houseman’s method^19^.

### Development of MPSs: methylation predictors of genetic principal components

We implemented a two-step feature selection method to identify the most representative CpG for each GPC in MPSs construction using training data. As shown in Figure 1, the first step involved a preliminary selection of CpG sites for each GPC based on their associations. As meta-analysis is shown to be equivalent to aggregated data analysis^23^, we estimated CpG site associations with GPCs by linear regression within each cohort’s training dataset in order to reduce computational burden. Each linear model is adjusted for age, sex, smoking and alcohol use status (if applicable), BMI, cell type proportions, study center, and race/ethnicity (if applicable; as identification with race/ethnicity group is often associated with lifestyle and environmental exposure patterns that may impact methylation). Specifically, smoking and alcohol use were modeled as categorical variable (0= never, 1=former or current). We then meta-analyzed these associations across cohorts by inverse variance, fixed effects meta-analysis. We applied False-Discovery Rate (FDR) correction on the resulting p-values using the Benjamini-Hochberg procedure^24^ and selected CpG sites with an FDR-adjusted q-value<0.05. We then aggregated the training datasets across all cohorts and applied weighted-Lasso regression to further refine the selection CpG sites and compute weights.

Weighted-Lasso was implemented as follows. We first constructed an initial MP (MPS_i_) for each GPC as weighted sums of the CpG sites identified by a first Lasso regression, adjusting for the same set of covariates (except for alcohol use due to partial availability) as were used in the initial regression analysis. Next, we added a weighting step because the variance of GPC varies by genetic ancestry due to patterns of allele frequency and linkage disequilibrium. Thus, each GPC was regressed on its corresponding, initial MPS_i_ without covariate adjustment to compute individual-specific weights, calculated as normalized squared residuals such that their sum equaled the sample size, with extreme values above 90^th^ percentile truncated to the 90^th^ percentile for stability^25^ (Figure 1). These individual weights were implemented as observation weights in a second Lasso regression to improve the accuracy of MPSs by accounting for differences in prediction variation related to genetic diversity (where genetic structure impacts genetic variance^26^).

### MPSs evaluation

We evaluated the performance of the MPSss, computed as weighted sums of CpG sites identified in the second Lasso regression, by computing their correlations with GPCs, and comparing them via visualization to GPCs and methylation PCs constructed via PCA based on CpG sites provided by Rahmani and colleagues^10^ and SNP adjacent CpG sites in the test dataset. Missing values in the methylation data were imputed using the mean across all samples before performing PCA. In particular, we performed visualization of population structure highlighting self-reported race/ethnicity groups, because race/ethnicity groups have shared patterns of genetic ancestry due to historical geographic migration patterns. We computed the variance explained by each MPSs for its corresponding GPC using linear regression adjusting for the same covariates aforementioned.

### Comparing alternative approaches to population stratification adjustment in epigenome-wide associations study of diabetes mellitus in HCHS/SOL

We also conducted epigenome wide association analysis (EWAS) in HCHS/SOL participants with diabetes as the exposure, which was defined according to medical history and lab criteria defined by American Diabetes Association, as previously described^27^. These analyses compared models that adjusted for MPSs in the subset of individuals who have genetic data and therefore have GPCs (n=1475), MPSs in a larger dataset of individuals who have methylation data (and therefore MPSs) but not necessarily GPCs (n=2695), models adjusted for GPCs alone (n=1475), and unadjusted models. In addition, we identified significant associations (adjusted p-value using Bonferroni correction less than 0.05) from the EWAS results in HCHS/SOL and compared them to previously identified diabetes associated CpG sites to evaluate biological relevance^28^. To further evaluate generalizability, we constructed MPSs in HCHS/SOL participants without genetic data and assessed their ability to differentiate individuals of different Hispanic/Latino backgrounds.

## Results

Descriptive statistics of demographic characteristics for the TOPMed cohorts, stratified by self-reported race/ethnicity, are summarized in Table 1. Across all cohorts, women comprised a higher proportion of participants. The average age ranges from 40 years in CARDIA to 60 years in MESA.

**Table 1.** Demographic characteristics of study cohorts.

| **Race/**<br>**Ethnicity** | **N** | **Age**<br>**(Mean (SD))** | **Sex**<br>**(N, %) - Female** | **BMI**<br>**(Mean (SD))** |
| --- | --- | --- | --- | --- |
| **ARIC** |  |  |  |  |
| White | 1461 | 52.34 (5.49) | 869 (59.5%) | 26.10 (4.19) |
| Black | 596 | 53.03 (5.62) | 366 (61.4%) | 29.80 (6.03) |
| **CARDIA** |  |  |  |  |
| White | 613 | 40.61 (3.40) | 366 (59.7%) | 27.13 (6.05) |
| Black | 510 | 39.68 (3.84) | 355 (69.6%) | 30.98 (7.70) |
| **JHS** |  |  |  |  |
| Black | 1365 | 55.89 (12.01) | 853 (62.5%) | 31.94 (7.36) |
| **MESA** |  |  |  |  |
| White | 396 | 60.91 (9.77) | 199 (50.3%) | 28.02 (4.81) |
| Black | 183 | 60.96 (9.57) | 106 (57.9%) | 30.64 (5.62) |
| Chinese | 69 | 60.97 (10.19) | 32 (46.4%) | 24.72 (3.05) |
| Hispanic/Latino | 281 | 58.65 (9.35) | 154 (54.8%) | 29.66 (5.03) |
| **HCHS/SOL** |  |  |  |  |
| Central American | 88 | 56.99 (7.87) | 58 (65.9%) | 30.57 (6.11) |
| Cuban | 359 | 57.39 (7.32) | 197 (54.9%) | 29.69 (5.31) |
| Dominican | 222 | 56.36 (7.87) | 164 (73.9%) | 29.92 (5.13) |
| Mexican | 298 | 55.88 (7.52) | 211 (70.8%) | 30.77 (5.92) |
| Puerto Rican | 412 | 57.75 (7.73) | 261 (63.3%) | 30.88 (6.25) |
| South American | 59 | 55.97 (6.42) | 43 (72.9%) | 29.54 (5.42) |
| Other/more than 1 | 36 | 57.44 (8.45) | 18 (50.0%) | 30.74 (5.69) |

MESA: Multi-Ethnic Study of Atherosclerosis; CARDIA: Coronary Artery Risk Development in Young Adults; JHS: Jackson Heart Study; ARIC: Atherosclerosis Risk in Communities study; HCHS/SOL: Hispanic Community Health Study/Study of Latinos

### Methylation principal components development

We developed MPSs for the first 10 GPCs. The number of CpG sites selected for each GPC in step 1 (FDR <0.05) and by the weighted Lasso regression in step 2 are reported in Table S1, with GPC1 having the largest number of associated CpGs (n= 32172), and GPC7 the fewest (n= 44). Detailed lists of CpGs selected for MPSs construction, along with weights are provided in the Supplementary Data 1. The top four PCs accounted for the majority of selected CpGs, whereas PC7 was associated with only 25 CpGs (Table S1).

### Methylation principal components evaluation

We constructed the MPSs in the aggregated TOPMed validation dataset (n=1090). Figure 2 visualizes the Pearson correlations between the 10 MPSs and GPCs. The MPSs were moderately to highly correlated with their corresponding GPCs. The strongest correlation was between the first MPSs and the first GPC (R^2^ = 0.98), with the lowest correlation between MPS7 and GPC7 (R^2^= 0.28). Similar to the correlation pattern among GPCs, MPS2 through MPS4 showed relatively higher correlations compared to other components. Interestingly, MPS5 and MPS8 demonstrated moderate correlations with GPC2 through GPC4 as well as their corresponding MPS—a pattern not observed for GPC5 and GPC8. Notably, the top three MPSs explained more than 80% of variance in their corresponding GPC, whereas the variance explained by MPS7 and MPS8 dropped below 10% (Table S2). The MPSs constructed using the 450K array follow a similar pattern, albeit with slightly less variance explained (Table S3).

**Figure 2.**
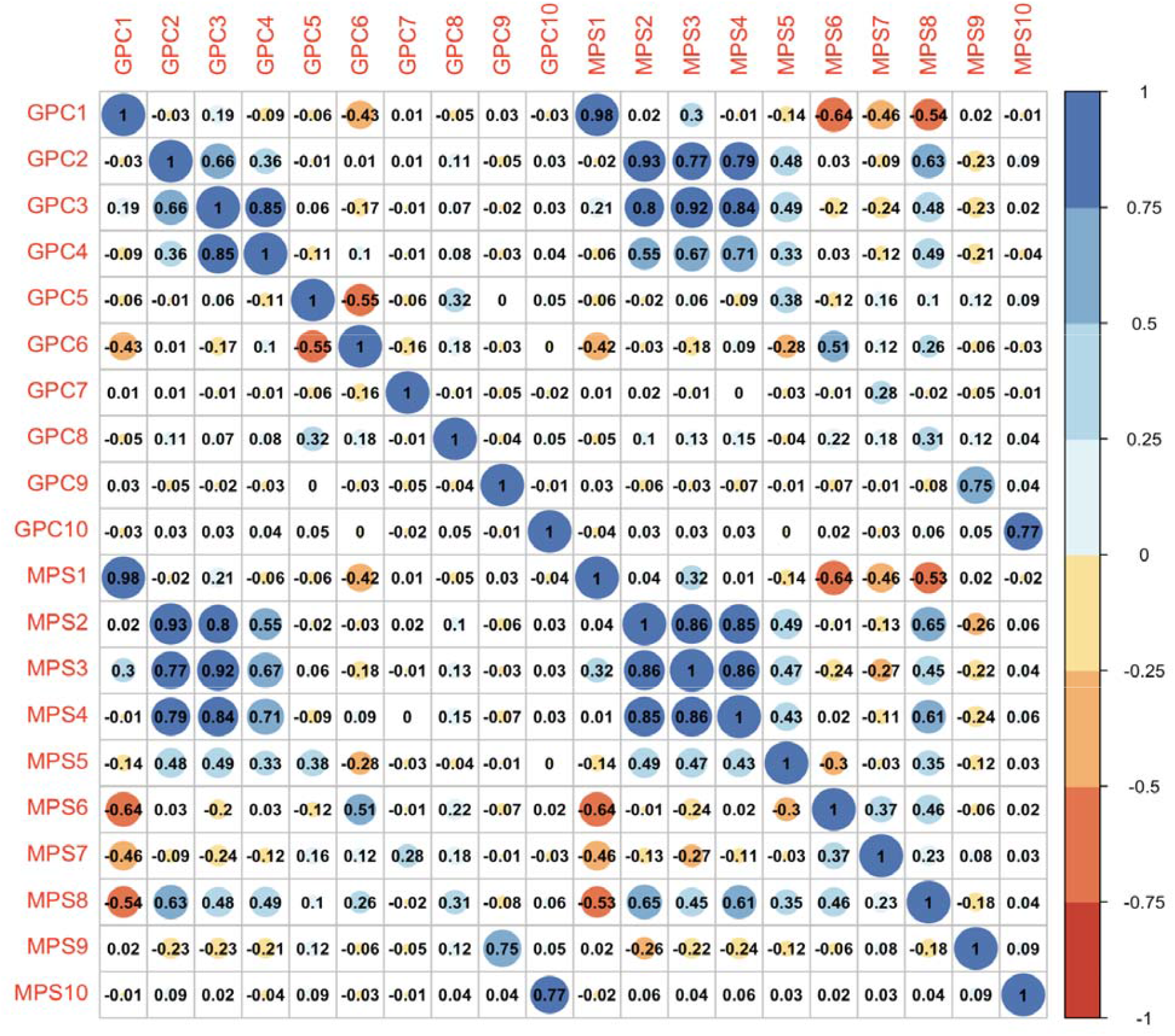
Correlation analysis between the 10 methylation (MPSs) and genetic principal components (PC).

Out of 4913 CpGs previously reported by Rahmani et al^10^. for construction of population stratification indicators, 1369 were available in our methylation data and used to construct the methylation principal components (MPC_Rahmani) in the aggregated test dataset across five TOPMed cohorts.

We visualized the top three principal components of our MPSs, MPC_Rahmani, MPC based on SNP adjacent CpG sites, and GPCs using scatterplots to assess their performance in differentiating the four self-reported race/ethnicity groups in our test dataset (Figure 3), which show group-level separation in GPCs space due to correlation of these groupings with genetic ancestry at the population level. We anticipated that methylation derived MPSs and MPCs showing similar patterns to GPCs. As shown in Figure 3A and 3D, MPSs derived from feature selection in this study exhibited clear separation among the four race/ethnicity groups, closely mirroring the clustering observed with GPCs, though the within-group dispersion appeared greater for MPSs. Additionally, the separation by MPSs between Chinese and Hispanic/Latino groups was less pronounced (Figure 3A). On the other hand, the top 3 MPC_Rahmani effectively distinguished Black, White and Hispanic/Latino participants, but not Chinese (Figure 3B). The clustering of the three race/ethnicity groups was more dispersed and less compact compared to MPSs (Figure 3A-3B). MPCs constructed using PCA on SNP adjacent CpG sites distinguishes Hispanic/Latino participants fairly well from other race/ethnicity groups, but Black and White participants are mixed together (Figure 3C). Moreover, a group of participants from all four race/ethnicity groups were separated out by MPCs (Figure 3C). Figure S1 illustrates the separation of self-reported Hispanic backgrounds in HCHS/SOL participants by MPSs, combining those in test data and without genetic data (n=689) (left), compared with the ones in the test dataset by GPCs (n=221) (right). While the self-reported Hispanic/Latino groups appear less distinct from each other in the MPSs space (left), it broadly replicates the structure observed with GPCs. In both cases, individuals of Mexican, Central American and South American backgrounds are more mixed together, while individuals of Dominican and Puerto Rican backgrounds were grouped closer to each other (Figure S1). According to the parallel coordinate plot, these backgrounds diverge more clearly at GPC3 and GPC8, as well as at MPS3 and MPS8 (Figure S2). However, Hispanic/Latino backgrounds exhibit minimal differentiation at MPS5 to MPS7 (Figure S2B).

**Figure 3.**
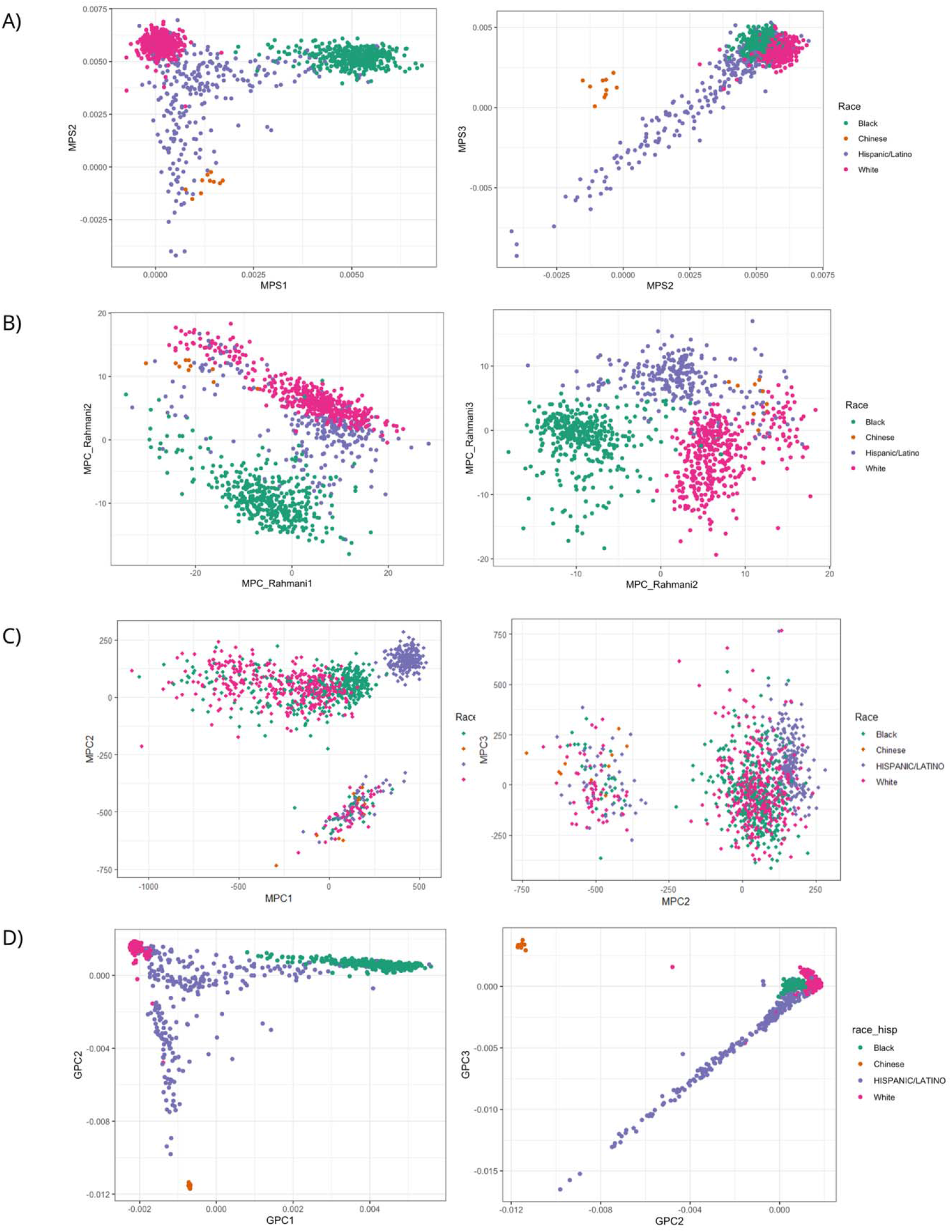
Scatter plots of GPCs, MPSs, and MPC_Rahmani, colored by race/ethnicity groups present in the data. A) Methylation population scores (MPSs) constructed in the current study; B) MPCs constructed via principal component analysis (PCA) using CpG sites from previously published paper by Rhamani et al.; C) Methylation PCs (MPC) constructed in test dataset through PCA on SNP adjacent CpGs; D) Genetic principal components (GPCs).

### Comparing population stratification adjustment in epigenome-wide association study of diabetes in HCHS/SOL

When using participants with genetic data (n= 1475), i.e. using exactly the same sample size to compare population stratification adjustment approaches, all three models appear inflated in both QQ plots and Manhattan plots (Figure 4a, Figure S2). The genomic inflation factors were 1.57 for no adjustment, 1.51 and 1.50 for adjusting for 5 GPCs and 5 MPSs, respectively, indicating moderate inflation across all three analyses, and some attenuation of inflation when adjusted for GPCs or MPSs. The number of statistically significant sites differed. Adjusting for 5 GPCs or 5 MPSs resulted in 153 and 157 significant CpGs, respectively, while no adjustment for population structure yielded 211 CpGs associated with diabetes (Figure S2). Figure 4B illustrates the results expanding the analysis to all available samples for no adjustment and adjusting for MPSs (n= 2695), with 2672 and 1749 significant CpGs, respectively. We also benchmarked the identified CpG sites against a previously validated set of 56 CpG sites associated with type 2 diabetes^28^. Of these sites, 15 have FDR p-value<0.05 (computed over the 56 sites) in the analysis that did not adjust for population structure and MPS-adjusted EWAS, and 14 for GPC-adjusted EWAS (Table S4). In the analysis using all available samples, 26 and 24 were identified for no-adjustment and MPS-adjusted EWAS, respectively (Table S5).

**Figure 4.**
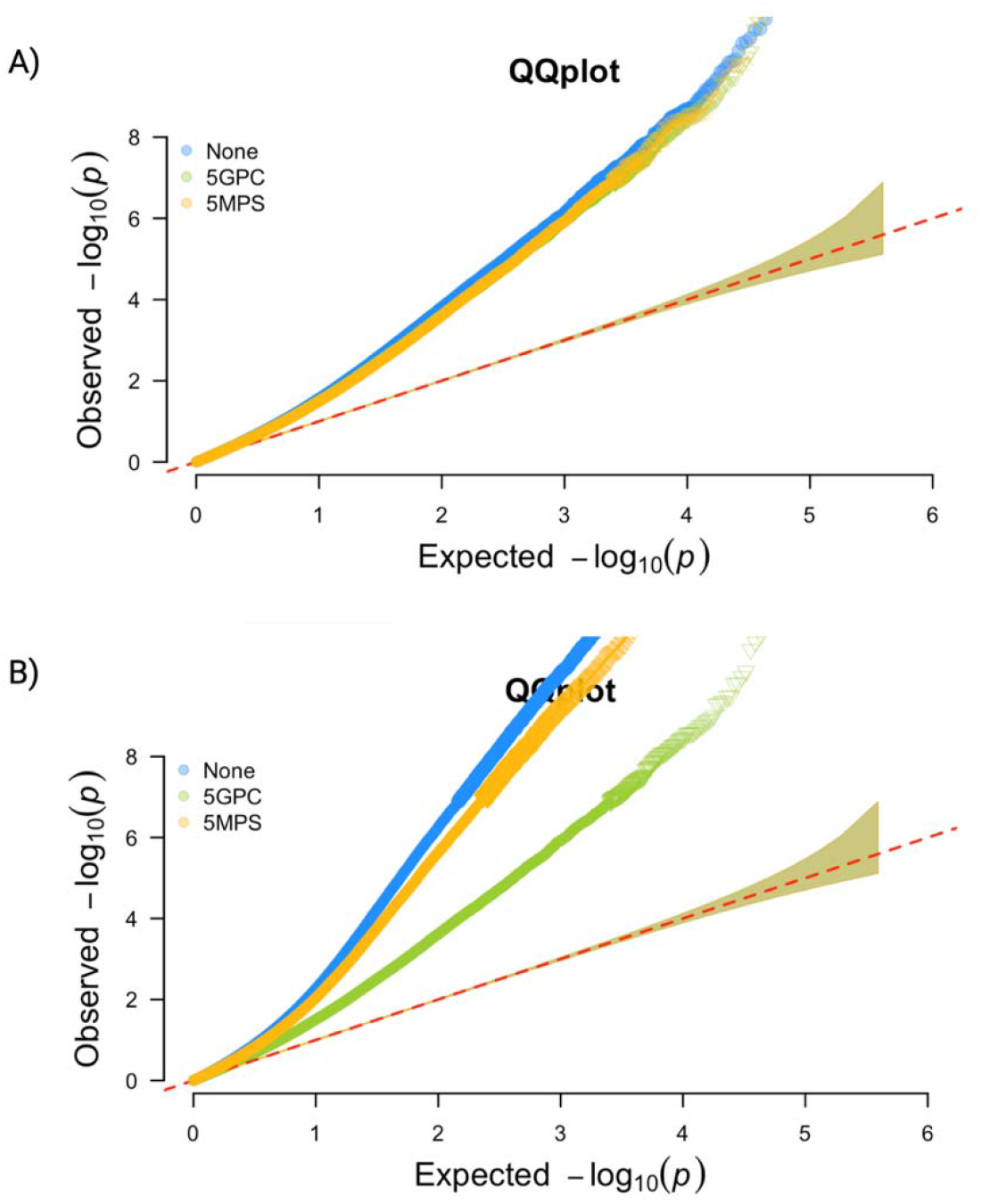
QQplot of results from EWAS adjusting for 5 genetic principal components (5GPCs), 5 methylation population scores (5MPSs) and neither (None). A) using same sample size (n=1475); B) using optimized sample size (n=2695 for MPSs adjusted model (5MPSs) and unadjusted model (None); n=1475 for GPC adjusted model)

## Discussion

We developed MPSs using DNAm data from multiple multi-ethnic cohorts via a two-step feature selection approach and evaluated their ability to capture population structure. The top three MPSs exhibited strong correlations with corresponding GPC and effectively separated self-reported race/ethnicity groups in the independent test dataset. On the other hand, MPCs derived from PCA using SNP adjacent CpG sites show inferior performance to MPSs in distinguishing across different race/ethnicity groups. The grouping of several individuals from distinct population backgrounds may be attributed to a certain degree of environmental effect captured by MPCs or the noise generated during the imputation step. While GPCs remain the gold standard to account for population structure in EWAS, our results indicate that MPSs offer comparable performance to GPCs and superior to unadjusted analyses, particularly when genetic data are unavailable. However, the moderate inflation factor suggests residual confounding that remains unaccounted for, likely attributable to unmeasured confounders or technical artifacts. This residual inflation underscores the importance of rigorous study design and analytical approaches to minimize confounding in EWAS^29,30^.

The distinction of self-reported Hispanic backgrounds by MPSs among HCHS/SOL participants without genetic data further supports their consistency in capturing meaningful population structure. The closer distance between Mexican, Central American, and South American Hispanic/Latino groups may be caused by more substantial proportions of Amerindian genetic ancestry compared to other Hispanic/Latino groups^31^. In contrast, the genetic ancestry of Dominican and Puerto Rican individuals is less well captured by the available CpG sites in the current dataset. This limitation could result from the pre-processing/quality control step of the methylation data, where some CpG sites were removed/masked in order to reduce bias.

Similar to the previously proposed methods to develop methylation-based variables that capture population structure by applying PCA over CpGs near SNPs^8,10,11^, our approach also relies on the assumption that some DNAm sites are associated with genetics^6,32^. Nevertheless, we select CpGs based on their associations with GPCs without assuming prior knowledge of ancestry-informative SNPs and flanking regions. Genetic PCs were treated as outcomes in our selection model to test the association between CpG sites and genetic PCs. While it is biologically reasonable to model CpG sites as outcomes, we retained genetic PCs as outcomes for consistency (Figure 1) due to single-outcome requirement for flexible feature selection method. Note that it is appropriate to use GPCs as outcomes and CpG as exposures because covariates effects are “regressed out” of the CpGs based on the Frisch–Waugh–Lovell theorem, just like covariates would be “regressed out” of the GPCs if they are used as exposures while CpGs are outcomes^33^. This method is also less computational demanding than traditional PCA when applied to large cohort data. Such efficiency was gained by first filtering out DNAm methylation sites not associated with genetic PCs, which can also be achieved by pre-selecting those associated with genetic variants. To further account for environmental confounders and tissue heterogeneity, factors previously observed to influence the selection of methylation sites in SNP-based methods^11^, we incorporated these variables as covariates in our models.

Nevertheless, the greater dispersion patterns of points within each group in the scatterplots (Figure 3) and lower correlations between non-top MPSs and GPCs imply that these MPSs do not entirely captured the genetic structure in the data. It is possible that heterogeneity in methylation data attributable to cohort-specific differences in quality control and data processing limited inference and performance of MPSs. Another limitation of this study is the small number of Chinese participants (n= 69, Table 1), who primarily have East Asian genetic ancestry, whereas TOPMed individuals self-reporting other race/ethnicities typically have low levels of East Asian genetic ancestry (See Supplementary Figure 3 in Kurniansyah et al. 2023^34^). This could partly explain their closer grouping towards Hispanics/Latinos participants, compared to when using GPCs (Figure 3). This impacts the generalizability of our MPSs in distinguishing the Chinese population in other independent datasets.

Both MPC_Rahmani derived from ancestry-informative SNPs and MPSs trained on GPCs effectively capture the prominent population structure. One limitation for the comparison with MPC created based on Rahmani et al.^10^ is the limited overlap between their selected CpG and those available in our integrated dataset (1469 out of 4913 CpGs). Moreover, their reference CpGs were selected using 450k DNAm data from individuals of European ancestry, which may restrict generalizability to other populations and to DNAm data generated using different arrays. Future research could therefore focus on enhancing the reproducibility and transferability of DNAm-based population structure prediction across diverse platforms and ancestries, e.g. by creating imputation models for missing CpG sites. To facilitate the use of our computed MPSs for future studies, we have made the lists of CpGs selected for each GPC, along with the corresponding R code, publicly available at Supplementary Data 1 and on Zenodo repository.

## Conclusion

Methylation-based scores predicting GPC, developed while integrating feature selection and adjustment for relevant confounders, provide a reliable estimate of population structure, showing strong concordance with GPCs, effective differentiation of racial and ethnic groups, as well as robust control of inflation in EWAS when genetic data are absent. With appropriate application, MPSs can complement GPCs to account for population structure in large cohorts when genetic data are unavailable for all or some individuals.

## Data Availability

TOPMed freeze 10 WGS, methylation, and phenotype data are available by application to dbGaP according to the study specific accession: ARIC: phs001211, CARDIA: phs001612, JHS: phs000964, HCHS/SOL: phs001395. JHS methylation used in this manuscript are available via application to dbGaP, via accession phs000286. JHS methylation data can also be accessed through data use agreement to coordinating center (https://www.jacksonheartstudy.org/). HCHS/SOL methylation data used in this manuscript are available through application to the database of Genotypes and Phenotypes (dbGaP) accession phs000810, or via data use agreement with the HCHS/SOL Data Coordinating Center (DCC) at the University of North Carolina at Chapel Hill, see collaborators website: https://sites.cscc.unc.edu/hchs/. MPSs CpGs and weights will be provided at the Zenodo repository.

## Acknowledgements

This work was supported by National Heart Lung and Blood Institute grant R01HL161012. Molecular data for the Trans-Omics in Precision Medicine (TOPMed) program was supported by the National Heart, Lung and Blood Institute (NHLBI). See the TOPMed Omics Support Table (Supplementary Note) for study specific omics support information. Core support including centralized genomic read mapping and genotype calling, along with variant quality metrics and filtering were provided by the TOPMed Informatics Research Center (3R01HL-117626-02S1; contract HHSN268201800002I). Core support including phenotype harmonization, data management, sample-identity QC, and general program coordination were provided by the TOPMed Data Coordinating Center (R01HL-120393; U01HL-120393; contract HHSN268201800001I). We gratefully acknowledge the studies and participants who provided biological samples and data for TOPMed. Study specific acknowledgements will be provided in the supplementary information.

