## Supplementary file for "Estimating population structure using epigenome-wide methylation data"

### Supplementary Tables

#### Table S1. CpG counts in methylation population score (MPS) construction from step 1 and step 2 (EPIC array)

| **Genetic principal components (GPC)** | **# CpG from Step 1** | **# CpG near SNPs (Step 1)** | **# CpG from Step 2** | **# CpG near SNPs (Step 2)** |
| --- | --- | --- | --- | --- |
| GPC 1 | 32172 | 26866 | 2218 | 1893 |
| GPC 2 | 15210 | 12772 | 2158 | 1835 |
| GPC 3 | 16249 | 13688 | 1674 | 1421 |
| GPC 4 | 11241 | 9445 | 1225 | 1053 |
| GPC 5 | 1381 | 1159 | 578 | 477 |
| GPC 6 | 356 | 306 | 175 | 152 |
| GPC 7 | 44 | 31 | 25 | 17 |
| GPC 8 | 10472 | 8708 | 690 | 574 |
| GPC 9 | 194 | 158 | 114 | 91 |
| GPC 10 | 131 | 111 | 87 | 73 |

Step 1 refers to regressions of individual CpG sites, where sites were selected if their FDR-corrected p-value in association with the corresponding GPC was < 0.05. Step 2 refers to the weighted LASSO of all CpG sites from step 1. CpG is considered to be near a SNP if it is up to 10 base-pairs away from a SNP, as annotated in the Illumina manifest of the chip.

SNP: single nucleotide polymorphism; LASSO: least absolute selection and shrinkage operator.

#### Table S2. Genetic PCs (GPCs) variance explained by methylation population scores (MPSs) using EPIC methylation array

| Principal component | Estimate | SE | P-value | Variance Explained |
| --- | --- | --- | --- | --- |
| GPC1 – MPS1 | 0.95 | 0.016 | < 10-300 | 96.98% |
| GPC2 – MPS2 | 1.05 | 0.012 | < 10-300 | 87.09% |
| GPC3 – MPS3 | 1.15 | 0.013 | < 10-300 | 84.86% |
| GPC4 – MPS4 | 1.07 | 0.020 | < 10-300 | 50.69% |
| GPC5 – MPS5 | 0.97 | 0.036 | 4.99E-123 | 13.60% |
| GPC6 – MPS6 | 0.80 | 0.058 | 1.04E-39 | 25.80% |
| GPC7 – MPS7 | 1.43 | 0.107 | 5.56E-38 | 8.17% |
| GPC8 – MPS8 | 1.03 | 0.040 | 3.02E-115 | 9.49% |
| GPC9 – MPS9 | 1.09 | 0.026 | 8.29E-232 | 56.01% |
| GPC10 – MPS10 | 1.11 | 0.027 | 8.41E-222 | 59.76% |

#### Table S3. Genetic PCs (GPCs) variance explained by methylation population scores (MPSs) constructed using 450K methylation array

| Principal component | Estimate | SE | P-value | Variance Explained |
| --- | --- | --- | --- | --- |
| GPC1 – MPS1 | 1.06 | 0.019 | 1.25E-312 | 96.26% |
| GPC2 – MPS2 | 1.06 | 0.015 | 0 | 84.83% |
| GPC3 – MPS3 | 1.24 | 0.016 | 0 | 77.87% |
| GPC4 – MPS4 | 1.41 | 0.030 | 6.40E-260 | 45.95% |
| GPC5 – MPS5 | 0.93 | 0.038 | 1.18E-105 | 13.93% |
| GPC6 – MPS6 | 0.95 | 0.067 | 2.89E-42 | 27.27% |
| GPC7 – MPS7 | 1.24 | 0.112 | 4.46E-27 | 4.26% |
| GPC8 – MPS8 | 1.01 | 0.048 | 1.05E-83 | 7.67% |
| GPC9 – MPS9 | 1.08 | 0.038 | 8.16E-133 | 38.65% |
| GPC10 – MPS10 | 1.09 | 0.042 | 7.51E-133 | 33.42% |

#### Table S4. CpGs associated with diabetes identified in EWAS in participants with genetic data

| CpGs | Gene | Model |
| --- | --- | --- |
| cg02050917 | SKI | None, 5GPC, 5MPS |
| cg05778424 | AKAP1 | None, 5GPC, 5MPS |
| cg06378491 | MAP4K2 | None, 5MPS |
| cg08309687 |  | None, 5GPC, 5MPS |
| cg08788930 | DENND3 | None, 5GPC, 5MPS |
| cg10639435 | ZNF250 | None, 5GPC, 5MPS |
| cg13059136 | SNORA54 | None, 5GPC, 5MPS |
| cg14020176 | SLC9A3R1 | None, 5GPC, 5MPS |
| cg14476101 | PHGDH | None, 5GPC, 5MPS |
| cg15020801 | PNPO | None, 5GPC, 5MPS |
| cg17901584 | DHCR24 | None, 5GPC, 5MPS |
| cg21480264 | POLN | None, 5GPC, 5MPS |
| cg24259291 | ZNFX1 | None, 5GPC, 5MPS |
| cg25316512 | ATN1 | None, 5GPC, 5MPS |
| cg27243685 | ABCG1 | None, 5GPC, 5MPS |

EWAS was performed in the HCHS/SOL. CpGs are from models using participants with genetic data (n= 1475), adjusted for 5 genetic principal components (GPC), 5 methylation population scores (MPS) and “none” (no adjustment to population structure)

#### Table S5. CpGs associated with diabetes identified in EWAS using all available individuals

| CpGs | Gene | Model |
| --- | --- | --- |
| cg01373896 | KLF16 | None |
| cg02050917 | SKI | None, 5MPS |
| cg04682775 | SLC6A9 | None, 5MPS |
| cg05778424 | AKAP1 | None, 5MPS |
| cg06378491 | MAP4K2 | None, 5MPS |
| cg06940720 |  | None, 5MPS |
| cg07719604 | ELMO3 | None, 5MPS |
| cg08309687 |  | None, 5MPS |
| cg08788930 | DENND3 | None, 5MPS |
| cg10192877 | ABCG1 | None |
| cg10639435 | ZNF250 | None, 5MPS |
| cg11202345 | LGALS3BP | None, 5MPS |
| cg13059136 | SNORA54 | None, 5MPS |
| cg14020176 | SLC9A3R1 | None, 5MPS |
| cg14476101 | PHGDH | None, 5MPS |
| cg15020801 | PNPO | None, 5MPS |
| cg17540192 | TECPR1 | None |
| cg17901584 | DHCR24 | None, 5MPS |
| cg18568872 | ZNF710 | None, 5MPS |
| cg19169154 | MFAP4 | None |
| cg21480264 | POLN | None, 5MPS |
| cg24259291 | ZNFX1 | None, 5MPS |
| cg24678869 | DENND4B | None, 5MPS |
| cg25178683 | LGALS3BP | None, 5MPS |
| cg25316512 | ATN1 | None, 5MPS |
| cg27243685 | ABCG1 | None, 5MPS |
| cg03691549 | TENC1 | 5MPS |
| cg14956201 | TRIO | 5MPS |

EWAS was performed in the HCHS/SOL. Models used all available samples (n=2695), and adjusted for 5 methylation population scores (MPS) and “none” (no adjustment to population structure).

### Supplementary Figures

#### Figure S1. Visualization of MPS by Hispanic/Latino background.

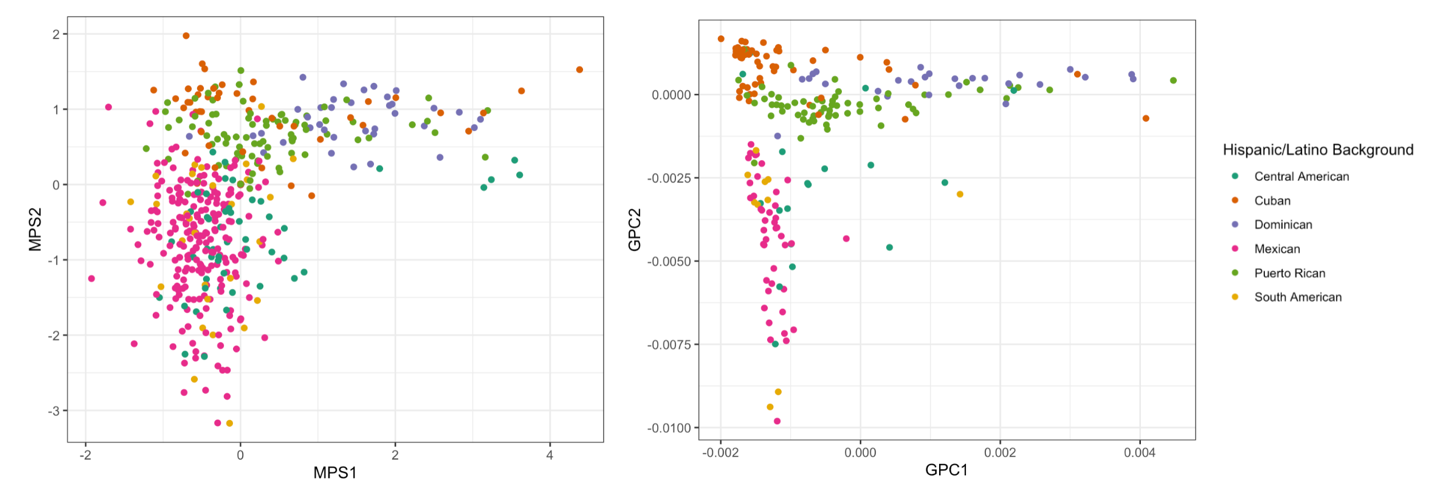

Figure S1. Scatter plots of methylation population scores (MPSs) constructed combining HCHS/SOL participants from test data and without genetic data (left) compared to genetic principal components (GPCs) of HCHS/SOL participants in test data (right) colored by Hispanic background.

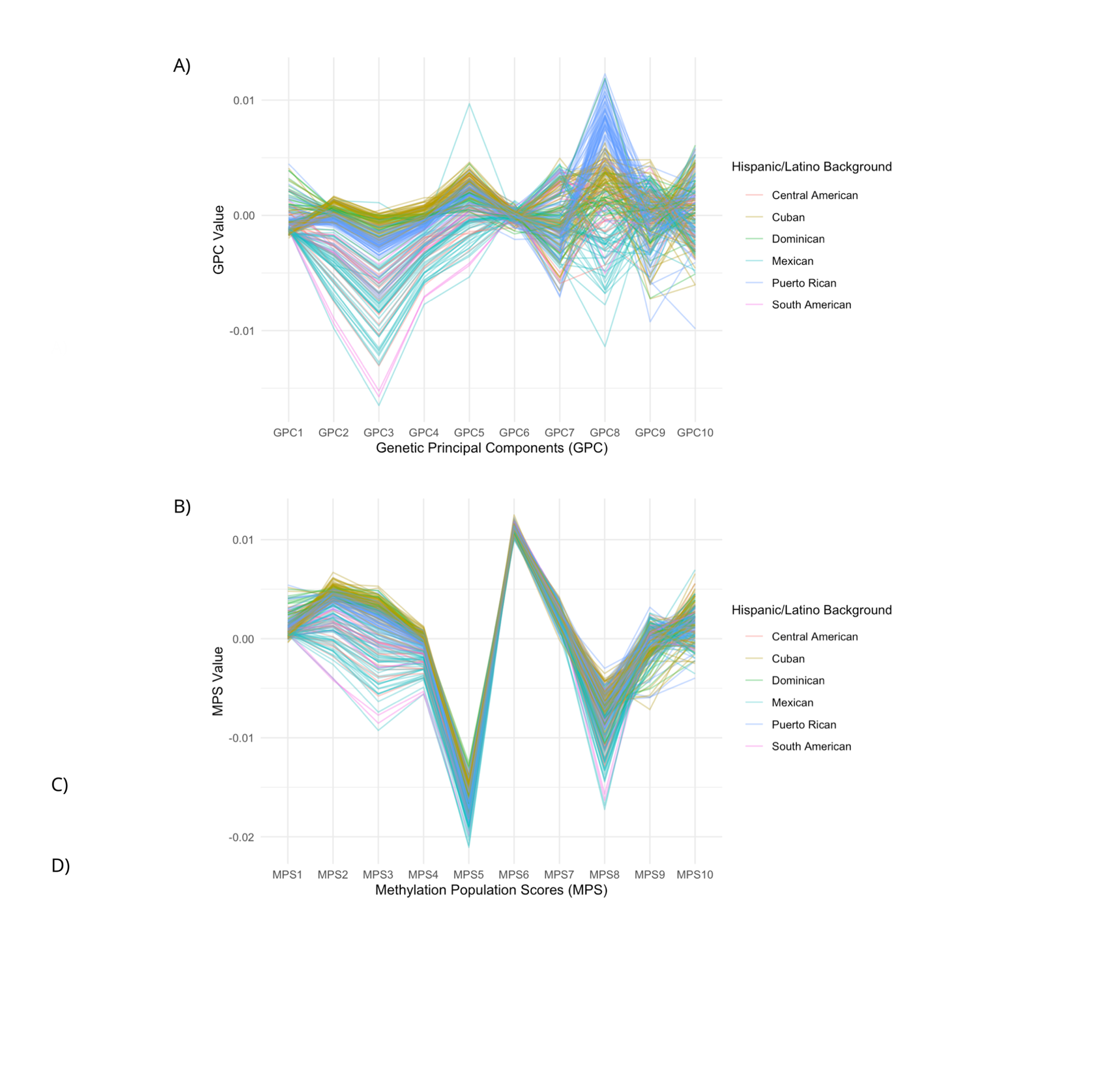

#### Figure S2. Parallel coordinate plots comparing GPSs and MPSs in HCHS/SOL.

Figure S2. Parallel coordinate plots for GPCs and MPSs across HCHS/SOL individuals, colored by Hispanic/Latino background

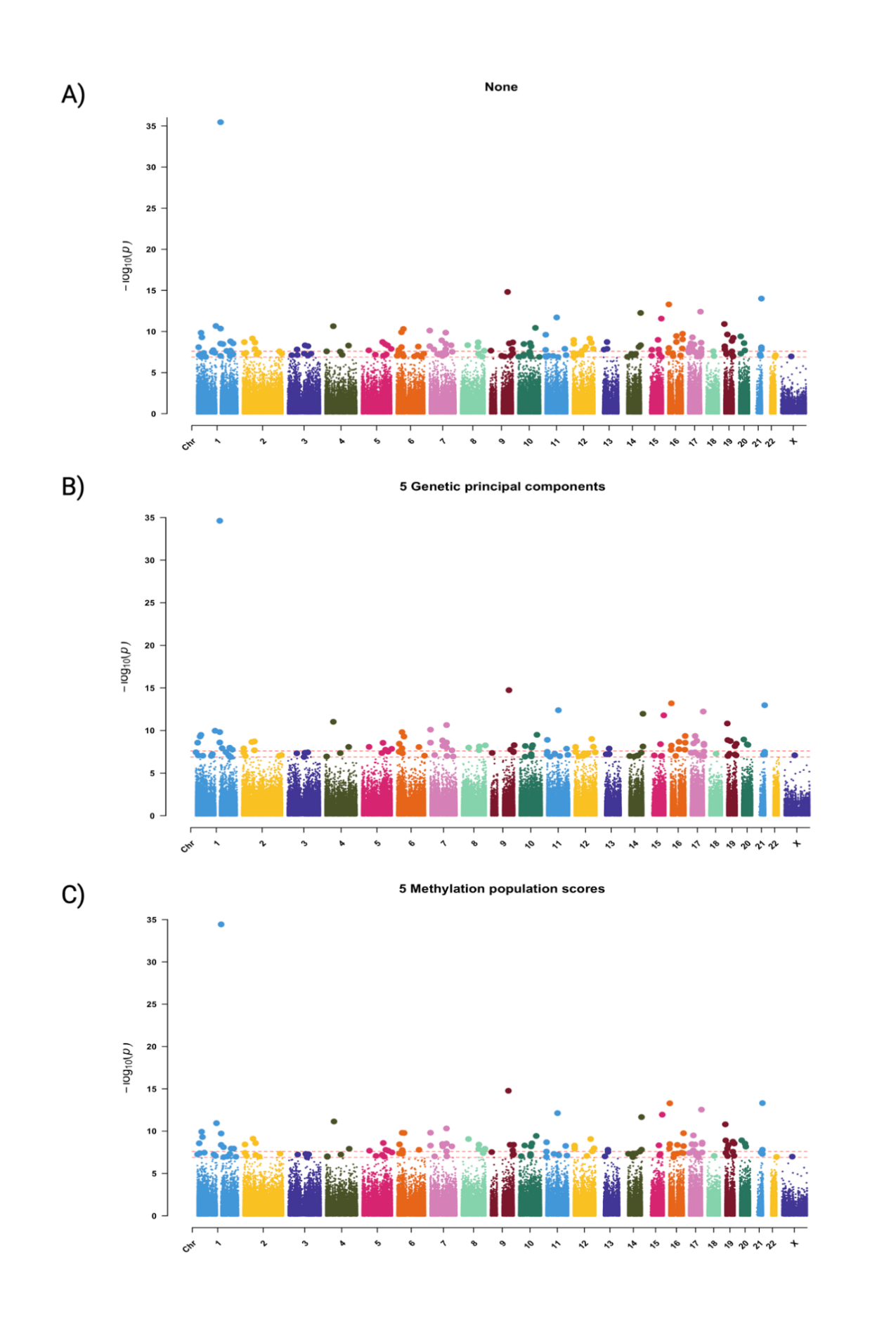

#### Figure S3. Manhattan plots of EWAS of diabetes in HCHS/SOL

Figure S3. Manhattan plots of epigenome-wide association analysis (EWAS) with diabetes as exposure (n=1475), of (A) unadjusted models; (B) adjusted for genetic principal components; (C) methylation population scores.

### Supplementary Note 1: TOPMed studies

#### The Atherosclerosis Risk in Communities (ARIC)

The ARIC Study is a biracial population-based longitudinal cohort study, recruiting 15,792 predominantly Black and White participants aged between 45 to 64 years by probability sampling from four U.S. communities, (Forsyth County, NC; Jackson, MS; suburban Minneapolis, MN, Minnesota; and Washington County, MD)^1^. Sociodemographic and lifestyle variables were collected through personal interviews and physical examinations at baseline (1987-1989) and multiple follow-up visits, including age, sex, race, smoking status, body mass index (BMI)^1^. The current study included 2057 ARIC participants (1,664 self-reported White and 674 self-reported Black) with genotype data that passed quality control from freeze 10.

**ARIC acknowledgements:**

Molecular data for the Trans-Omics in Precision Medicine (TOPMed) program was supported by the National Heart, Lung and Blood Institute (NHLBI). Genome sequencing for “NHLBI TOPMed – NHGRI CCDG: Atherosclerosis Risk in Communities (ARIC)” (phs001211) was performed at the Baylor College of Medicine Human Genome Sequencing Center (HHSN268201500015C and 3U54HG003273-12S2) and the Broad Institute for MIT and Harvard (3R01HL092577- 06S1). Methylation for “NHLBI TOPMed – NHGRI CCDG: Atherosclerosis Risk in Communities (ARIC)” (phs001211) was performed at the University of Washington Northwest Genomics Center (HHSN268201600032l). Core support including centralized genomic read mapping and genotype calling, along with variant quality metrics and filtering were provided by the TOPMed Informatics Research Center (3R01HL-117626-02S1; contract HHSN268201800002l). Core support including phenotype harmonization, data management, sample-identity QC, and general program coordination, were provided by the TOPMed Data Coordinating Center (R01HL-120393; U01HL-120393; contract HHSN268201800001l). We gratefully acknowledge the studies and participants who provided biological samples and data for TOPMed.

The Genome Sequencing Program (GSP) was funded by the National Human Genome Research Institute (NHGRI), the National Heart, Lung, and Blood Institute (NHLBI), and the National Eye Institute (NEI). The GSP Coordinating Center (U24 HG008956) contributed to cross program scientific initiatives and provided logistical and general study coordination. The Centers for Common Disease Genomics (CCDG) program was supported by NHGRI and NHLBI, and whole genome sequencing was performed at the Baylor College of Medicine Human Genome Sequencing Center (UM1 HG008898).

The Atherosclerosis Risk in Communities study has been funded in whole or in part with Federal funds from the National Heart, Lung, and Blood Institute, National Institutes of Health, Department of Health and Human Services under Contract nos. (75N92022D00001, 75N92022D00002, 75N92022D00003, 75N92022D00004, 75N92022D00005). The authors thank the staff and participants of the ARIC study for their important contributions.

#### The Jackson Heart Study (JHS)

The Jackson Heart Study is a community-based, observational study consisting of 5,306 non-institutionalized African-American participants aged between 35-84 years recruited from Jackson, MS, metropolitan statistical area (Hinds, Madison, and Rankin)^2^ during 2000–2004. Sociodemographic and lifestyle variables including family medical history were collected using self-reported questionnaires during clinic visit^3^. To avoid duplicated individuals across ARIC and JHS, we filtered out participants that were also sampled by the ARIC study, resulting in 1365 JHS participants in total.

**JHS acknowledgements:**

The Jackson Heart Study is supported by Contracts HHSN268201800010I, HHSN268201800011I, HHSN268201800012I, HHSN268201800013I, HHSN268201800014I, HHSN268201800015I from the National Heart Lung and Blood Institute (NHLBI) with additional support from the National Institute of Minority Health and Health Disparities (NIMHD).

The views expressed in this manuscript are those of the authors and do not necessarily represent the views of the National Heart, Lung, and Blood Institute; the National Institute of Minority Health and Health Disparities (NIMHD); the National Institutes of Health; or the U.S. Department of Health and Human Services. Genome sequencing (dbGap accession phs000964) was performed at the Northwest Genomics Center (HHSN268201100037C). Core support including centralized genomic read mapping and genotype calling, along with variant quality metrics and filtering were provided by the TOPMed Informatics Research Center (R01HL117626; contract HHSN268201800002I). Core support including phenotype harmonization, data management, sample-identity QC, and general program coordination were provided by the TOPMed Data Coordinating Center (R01HL120393; U01HL120393; contract HHSN268201800001I).

#### The Multi-Ethnic Study of Atherosclerosis (MESA)

MESA is a multi-ethnic longitudinal cohort study composed of 6,814 participants aged 45–84 years from six field centers (Baltimore, MD; Chicago, IL; Forsyth County, NC; [Los Angeles](https://www.sciencedirect.com/topics/earth-and-planetary-sciences/los-angeles), CA; New York, NY; and St Paul, MN), all free of overt clinical cardiovascular diseases at baseline (Exam 1; 2000-2002)^4^. This study included 929 participants from TOPMed MESA Multi-Omics project, with available genetic and methylation data collected from baseline, during which sociodemographic characteristics, such as age, sex, and race (White, African American, Hispanic/Latino, and Asian), were self-reported using standard questionnaires.

**MESA acknowledgements:**

Whole genome sequencing (WGS) and methylation data for the Trans-Omics in Precision Medicine (TOPMed) program was supported by the National Heart, Lung and Blood Institute (NHLBI). WGS for “NHLBI TOPMed: Multi-Ethnic Study of Atherosclerosis (MESA)” (phs001416.v1.p1) was performed at the Broad Institute of MIT and Harvard (3U54HG003067-13S1). Centralized read mapping and genotype calling, along with variant quality metrics and filtering were provided by the TOPMed Informatics Research Center (3R01HL-117626-02S1). Phenotype harmonization, data management, sample-identity QC, and general study coordination, were provided by the TOPMed Data Coordinating Center (3R01HL-120393-02S1). Methylation data for “NHLBI TOPMed: Multi-Ethnic Study of Atherosclerosis (MESA)” (phs001416) was performed at Keck Molecular Genomics Core Facility (HHSN268201600034I)."

The MESA projects are conducted and supported by the National Heart, Lung, and Blood Institute (NHLBI) in collaboration with MESA investigators. Support for MESA is provided by contracts 75N92025D00022, 75N92020D00001, HHSN268201500003I, N01-HC-95159, 75N92025D00026, 75N92020D00005, N01-HC-95160, 75N92020D00002, N01-HC-95161, 75N92025D00024, 75N92020D00003, N01-HC-95162, 75N92025D00027, 75N92020D00006, N01-HC-95163, 75N92025D00025, 75N92020D00004, N01-HC-95164, 75N92025D00028, 75N92020D00007, N01-HC-95165, N01-HC-95166, N01-HC-95167, N01-HC-95168, N01-HC-95169, UL1-TR-000040, UL1-TR-001079, UL1-TR-001420, UL1TR001881, DK063491, and R01HL105756.  The authors thank the MESA participants and the MESA investigators and staff for their valuable contributions.  A full list of participating MESA investigators and institutions can be found at [http://www.mesa-nhlbi.org](https://urldefense.com/v3/__http:/www.mesa-nhlbi.org__;!!AIv8Mrc!63FBlor7CCnRNhn6nLy7ZSgIqfMbe37c_sOtRRIfPrTvrOK01Y4qbO4MzfVFd2Bn_Ay8StYieBUg0Z-LI8yd1OxfXhSv$).

#### The Hispanic Community Health Study/Study of Latinos (HCHS/SOL)

The HCHS/SOL is a population-based cohort study, which recruited 16,415 men and women representing diverse Hispanic/Latino origins from four urban areas across the United States (Bronx NY, Chicago IL, Miami FL, and San Diego CA), with a wide age range from 18 to 74 years old^5^. Sociodemographic and lifestyle variables were self-reported at baseline exam during 2008-2011, including age, sex, smoking status, alcohol consumption, as well as Hispanic background, while BMI was measured following standard procedure. Our study consisted of 1949 HCHS/SOL participants, including 474 participants without genetic data.

**HCHS/SOL acknowledgements:**

The Hispanic Community Health Study/Study of Latinos is a collaborative study supported by contracts from the National Heart, Lung, and Blood Institute (NHLBI) to the University of North Carolina (HHSN268201300001I / N01-HC-65233), University of Miami (HHSN268201300004I / N01-HC- 65234), Albert Einstein College of Medicine (HHSN268201300002I / N01-HC-65235), University of Illinois at Chicago – HHSN268201300003I / N01- HC-65236 Northwestern Univ), and San Diego State University (HHSN268201300005I / N01-HC-65237). The following Institutes/Centers/Offices have contributed to the HCHS/SOL through a transfer of funds to the NHLBI: National Institute on Minority Health and Health Disparities, National Institute on Deafness and Other Communication Disorders, National Institute of Dental and Craniofacial Research, National Institute of Diabetes and Digestive and Kidney Diseases, National Institute of Neurological Disorders and Stroke, NIH Institution-Office of Dietary Supplements.

#### The Coronary Artery Risk Development in Young Adults study (CARDIA)

The CARDIA study is a multi-center population study of cardiovascular disease in 5,115 African Americans and whites aged between 18-30 years at the time of enrollment^6^. Specifically, the participants were recruited with proportionate racial, gender, age, and education groups from four U.S urban areas (Birmingham, AL; Chicago, IL; Minneapolis, MN; and Oakland, CA)^6^. Demographic variables such as age, race, and sex were self-reported and verified during the baseline clinic visit in 1985-1986, while DNA extraction for methylation profiling was conducted at year 15 and year 20 follow-up examinations^7,8^. Our study included 1123 CARDIA participants having both genetic and methylation data from year 15.

**CARDIA acknowledgements:**

The Coronary Artery Risk Development in Young Adults Study (CARDIA) is conducted and supported by the National Heart, Lung, and Blood Institute (NHLBI) in collaboration with the University of Alabama at Birmingham (75N92023D00002 & 75N92023D00005), Northwestern University (75N92023D00004), University of Minnesota (75N92023D00006), and Kaiser Foundation Research Institute (75N92023D00003). CARDIA was also partially supported by the Intramural Research Program of the National Institute on Aging (NIA) and an intra‐agency agreement between NIA and NHLBI (AG0005). The DNA methylation profiling and data processing were funded by the American Heart Association (17SFRN33700278 and 14SFRN20790000, Northwestern University, to Dr. Hou) and NHLBI TOPMed (MPIs: Drs. Hou and Lloyd-Jones).
